# Burnout among labor and birth providers in northern Tanzania: A mixed-method study

**DOI:** 10.1101/2023.05.28.23290395

**Authors:** Virginie Marchand, Melissa H. Watt, Linda M. Minja, Mariam L. Barabara, Janeth Mlay, Maya J. Stephens, Olivia Hanson, Blandina T. Mmbaga, Susanna R. Cohen

## Abstract

Burnout, characterized by emotional exhaustion, depersonalization, and a diminished sense of accomplishment, is a serious problem among healthcare workers. Burnout negatively impacts provider well-being, patient outcomes, and healthcare systems globally, and is especially worrisome in settings with a shortage of healthcare workers and resources. The goal of this study is to explore the experience of burnout in a population of labor and delivery (L&D) providers in Tanzania. We examined burnout using three data sources. A structured assessment of burnout was collected at four time points from a sample of 60 L&D providers in six clinics. The same providers participated in an interactive group activity from which we drew observational data on burnout prevalence. Finally, we conducted in-depth interviews (IDIs) with a subset of 15 providers to further explore their experience of burnout. At baseline, prior to any introduction to the concept, 18% of respondents met criteria for burnout. Immediately after a discussion and activity on burnout, 62% of providers met criteria. One- and three-months later, 29% and 33% of providers met criteria, respectively. In IDIs, participants saw the lack of understanding of burnout as the cause for low baseline rates and attributed the subsequent decrease in burnout to newly acquired coping strategies. The activity helped providers realize they were not alone in their experience of burnout. High patient load, low staffing, limited resources, and low pay emerged as contributing factors. Burnout was prevalent among a sample of L&D providers in northern Tanzania. However, a lack of exposure to the concept of burnout leads to providers being unaware of the issue as a collective burden. Therefore, burnout remains rarely discussed and not addressed, thus continuing to impact provider and patient health. Previously validated burnout measures cannot adequately assess burnout without a discussion of the context.

## INTRODUCTION

The COVID-19 pandemic focused international attention on healthcare provider burnout (1,2), but it has been a chronic problem in the healthcare setting due to high levels of stress and emotional intensity associated with the job (3). Provider burnout is characterized by three components: 1) emotional exhaustion, defined as the feeling of being “used up” and unavailable emotionally for patients at the end of the workday; 2) depersonalization, or increased callousness towards patients; and 3) a sense of diminished personal accomplishment, including feelings of ineffectiveness and lack of value to patient care (3).

Rates of burnout are higher among healthcare trainees and professionals compared to other professions (3,4). Burnout among healthcare personnel negatively impacts provider health and well-being, contributing to an increased risk of depression (3,5), alcohol misuse (6) and suicidal ideation (7). Provider burnout also has a significant impact on patient care and the healthcare system as a whole (3), with data showing a greater rate of medical errors (8), decreased productivity (9), increased desire to quit the profession (10), and diminished rapport with patients (11).

Burnout in low and middle income countries (LMICs), especially in Sub-Saharan Africa has been underexplored (12,13) and specific measures to assess provider burnout have not been validated. A systematic review of burnout among healthcare providers in Africa found that burnout was associated with a heavy workload, difficult work conditions, inadequate personnel, and low work satisfaction (12). A survey of burnout among East African nurses found a high rate of emotional exhaustion and depersonalization (14). In LMICs, the shortage of healthcare workers puts additional pressure on health systems (15); in Sub-Saharan Africa, nearly one fourth of the global disease burden is met by only 3% of the global work force (16).

Labor and delivery (L&D) wards are particularly high-stress environments, and research suggests that maternal health staff may be at higher risk of burnout compared to colleagues in other specialties (17). Given the impact of provider burnout on patient care, this could have important implications for maternal and child health outcomes.

While healthcare provider burnout has been part of the national conversation in the United States, and particularly among health care providers themselves, in the previous three years, it has not received the same level of attention in LMICs. The goal of this study was to comprehensively explore the understanding of burnout in a population of L&D providers in Tanzania, and how providers experience burnout in their work. We draw on structured assessment of burnout at four time points, observations during an interactive group activity on burnout, and semi-structured interviews. The findings shed light on the experience of burnout in Tanzania and can help provide guidance on measurement and intervention.

## METHODOLOGY

### Overview

As part of a larger study, our team engaged with providers, patients and stakeholders to develop, deliver and evaluate the MAMA simulation and team-based training, focused on respectful maternity care (RMC) during L&D for women living with HIV (WLHIV) in Tanzania (18). The 2.5-day training included case-based learning sessions, simulations, and interactive activities on teamwork and communication, clinical empathy, stigma and bias, and RMC. Further intervention content has been described elsewhere (18).

In developing the intervention, we conducted focus groups with providers and uncovered that burnout was a possible contributor to suboptimal delivery of RMC and an actionable area for intervention. High workloads and limited staffing in participating clinics led to providers feeling overworked and exhausted. However, these feelings were rarely discussed among providers. Therefore, we decided to incorporated assessments of, and content material on, burnout throughout the training. Data from these focus group discussions will be presented elsewhere. **Data Sources**

Study participants were 60 L&D providers from six primary care hospitals in the Moshi (urban) and Rombo (rural) districts of the Kilimanjaro region of northern Tanzania. The sample size of 60 was based on consultation with the Tanzania Ministry of Health, who felt that ten providers per clinic would provide adequate intervention coverage while not disrupting clinical flow. After obtaining written informed consents, participants were assigned a study ID. Assessments were conducted at four time points: baseline, immediate post, 1 month post, and 3 months post. The participant flow and follow-up rate is visualized **Supplement Table 1.**

We assessed burnout through three sources of data: observational data from an interactive activity during a training session on burnout, structured assessments of burnout through surveys, and in-depth interviews on burnout.

### Interactive session

The MAMA training was delivered to providers in November 2022. The curriculum included one 45-minute session focused specifically on burnout, followed by three sessions on clinical empathy (45 minutes), bias and stigma (45 minutes), and mindfulness and coping strategies (30 minutes). During the burnout session, we first defined burnout for participants as a combination of emotional exhaustion, depersonalization, and diminution of personal accomplishment, explaining this could present as feelings of emptiness, fatigue, negative or distant attitudes towards others, feelings of failure, or low self-esteem. We prompted providers to share their thoughts on factors contributing to burnout in their clinical settings and to engage in a discussion based on their experience. Then, we invited them to participate in an activity in which they lined-up in the room along a scale from “not burned out at all” to the left to “completely burned out” to the right to create a visual representation of the prevalence of burnout among this group of providers.

After the activity, we held a discussion of personal strategies to cope with stress in their high-pressure work environment including mindfulness and clinical empathy skills. These included deep breathing, body scan, positive affirmations, holding a powerful pose, daily intentions, stretching, writing down accomplishments, focusing on sensations during small activities like handwashing, taking a moment to pause and reflect on next steps when possible. Participants had the chance to practice each one together and we encouraged them to choose one or two to practice at work.

Additionally, the topics burnout, mindfulness and empathy were integrated throughout the rest of the MAMA trainings, which was designed to support providers’ emotional wellbeing and help them connect with themselves, their colleagues and patients through teamwork-based simulations, games, and opportunities for discussion and reflection.

### Survey Assessment

We assessed provider burnout using a 2-item measure (19) adapted from the Maslach Burnout Inventory (20), which were translated, back translated, and piloted with team members and a small group of providers to ensure accurate translation. After giving a written definition of burnout (Box 1), translated as “msongomoto” in Swahili, the first question asks respondent to select one of five responses: “1) I enjoy my work. I have no symptoms of burnout”; “2) Occasionally I am under stress, and I don’t always have as much energy as I once did, but I don’t feel burned out”; “3) I am definitely burning out and have one or more symptoms of burnout, such as physical and emotional exhaustion”; “4) the symptoms of burnout that I’m experiencing won’t go away. I think about frustrations at work a lot”; and “5) I feel completely burned out and often wonder if I can go on. I am at the point where I may need some changes or may need to seek some sort of help.” This question was scored on a 5-point Likert scale (0-4).

#### Box 1

##### Definition of burnout given to participants as part of the measure

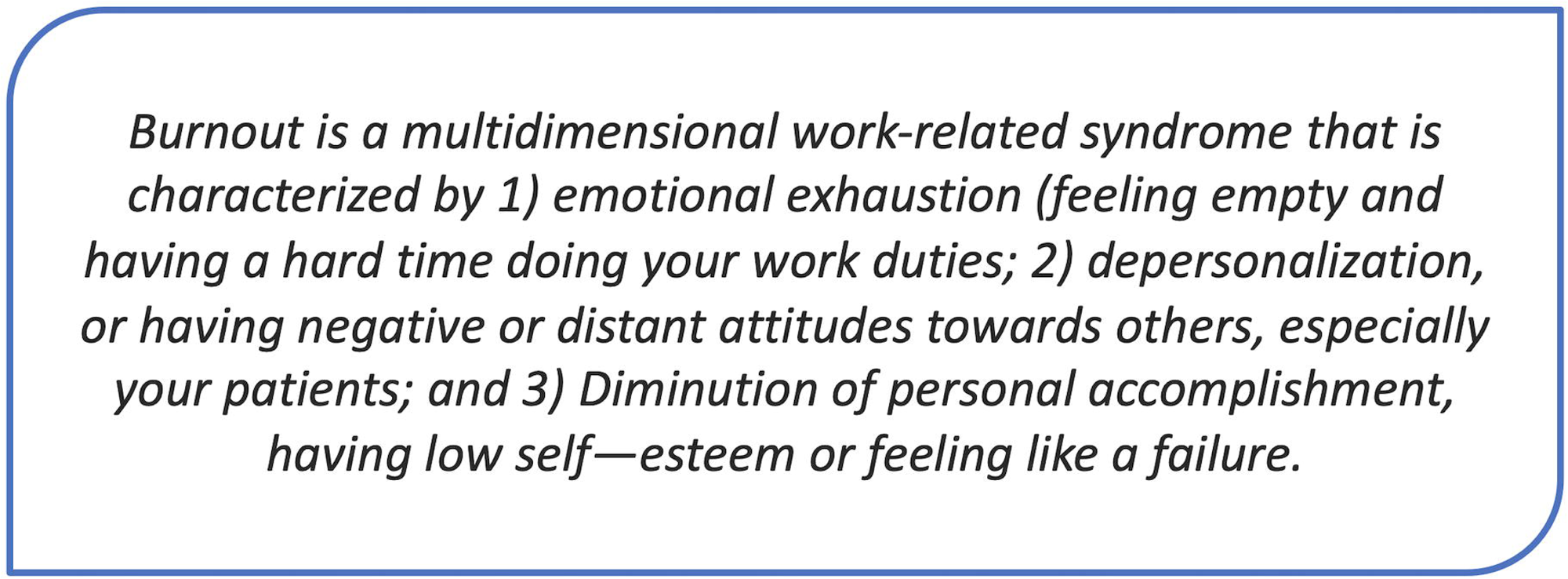

The second question asked participants how often they feel they have become more callous toward patients since taking this job because of burnout. The response was given on a scale of “Never” to “Every Day” and scored on a 7-point Likert scale (0-6).

We used this measure to assess burnout at four timepoints: immediately before the delivery of the MAMA training, immediately after the burnout session and interactive activity during the training, then again one- and three-months post-training. Using >2 as the cutoff for burnout for the first question (on a 0-4 scale) and >4 for the second question (on a 0-6 scale), we considered providers burned out if they met criteria for burnout in either question and classified providers categorically as positive or negative for burnout for our analysis (19,21).

### In-depth interviews

To further understand the experience of burnout in their clinical context, we invited a subset of 15 providers (2-3 from each facility) to further discuss burnout through in-depth interviews (IDIs) three to four months post training. We selected participants who had actively participated and shared their opinions and experiences during the MAMA training and were viewed by the team as “good informants” (22). IDI participants were further selected to ensure representation of gender, age, and profession. All 15 providers invited to participate in IDIs agreed. IDIs were conducted in Swahili by four research team members. During the IDIs, we asked providers if they felt that burnout was common among their colleagues and what it looks like. We asked if burnout was discussed among colleagues, both before and after the training, and if so, how it is described. We also explored the impact the training had on the way they view burnout in themselves and among colleagues. Finally, we applied the method of member checking by presenting interviewees with our preliminary survey data on reported burnout and asking for their interpretation of the trends between time-points based on their experience. This was the opportunity to ask them why they thought the reported rates of burnout on the initial surveys seemed lower than the levels of burnout expressed through conversation and displayed through the activity.

### Data analysis

Survey data were entered into REDCap software and exported to R for data analysis. We first used summary statistics to describe participant characteristics. We then determined the proportion of patients meeting criteria for burnout at each timepoint and used a Pearson’s Chi Squared test to assess changes overall and from mid-training to one-month post training. IDIs were transcribed and translated then exported into NVivo for applied thematic analysis (23). The data were coded to identify emerging themes across five domains: prevalence of burnout, drivers of burnout, manifestations of burnout, explanations of low baseline scores, and explanations for burnout changes over time. The research team met to discuss the codes in order to group and synthesize concepts using axial coding (23,24). Analysis was iterative throughout the data collection process to assess for thematic saturation (25), which was determined when no new themes or information emerged, and themes were felt to adequately describe the domain of interest.

### Ethical considerations

Written informed consent for participation in the MAMA training, completion of surveys and participation in interviews was obtained from all participants prior to initiation of the study. All participant information was stored in a locked cabinet and secured database. The study was approved by the ethical review committees at the University of Utah (Protocol 00143918), Kilimanjaro Christian Medical Center (Protocol 2056), and National Institute for Medical Research in Tanzania (Protocol 3853). The trial is registered at clinicaltrials.gov (NCT05271903).

## RESULTS

### Sample description

A total of 60 providers completed the interactive activity in two cohorts of thirty participants. Surveys were completed by sixty at timepoint 1 (pre-training), sixty at timepoint 2 (mid-training), fifty-five at timepoint 3 (one-month post-training), and fifty-nine at timepoint 4 (three-months post-training). Fifteen participants participated in the IDIs. Table 1 describes baseline characteristics of our sample of participating providers.

**Table 1:**
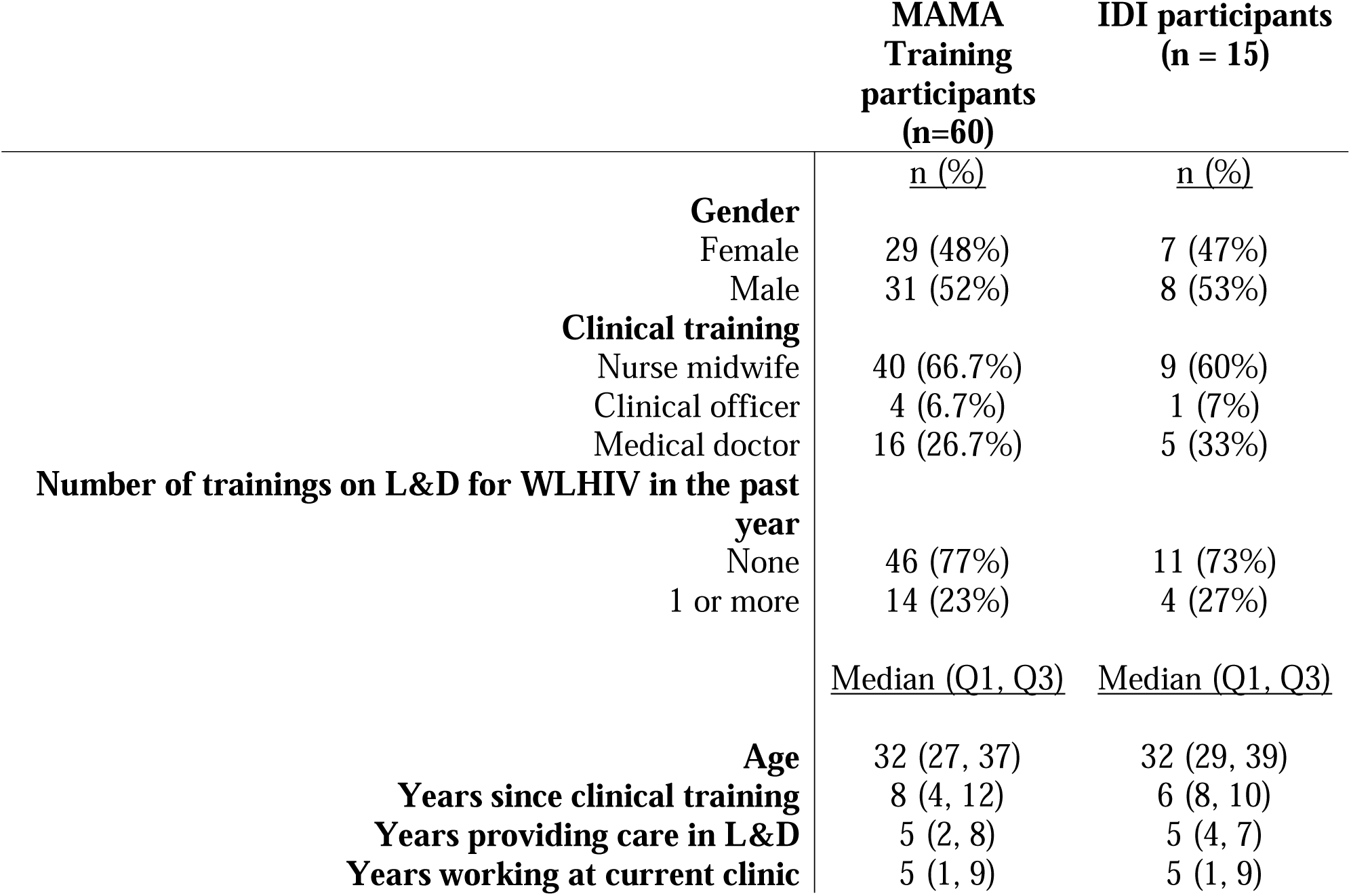
Description of the sample of L&D providers participating in the MAMA intervention

### Baseline assessment of burnout

In the baseline surveys prior to the training, only 11 providers (18%) met criteria for burnout. This came to the attention of the lead trainers who were curious about this number given the findings from our pre-training development phase, in which focus groups identified high stress, workload, and limited personnel as issues, all of which in the literature are linked to high burnout. This observation triggered the training team to make an on-the-spot alternation to the curriculum and to take the time to discuss the definition of burnout and explore the topic with participants through the interactive activity described below.

### Training session & burnout activity

During the large discussion of causes of burnout during the training, providers mentioned high patient load, low staffing, and limited resources. The robust discussion allowed for self-reflection and for providers to hear from their peers the range of experiences around burnout and resilience. Providers with more years of experience shared their coping mechanisms, and newer providers were able to air their frustrations and growing sense of overwhelm. At the conclusion of the discussion, providers were asked to physically place themselves along a continuum, from not at all burned out to completely burned out. The human scale revealed that a majority of participants felt burnt out, standing close together towards the right side of the scale (Figure 1).

**Figure 1:**
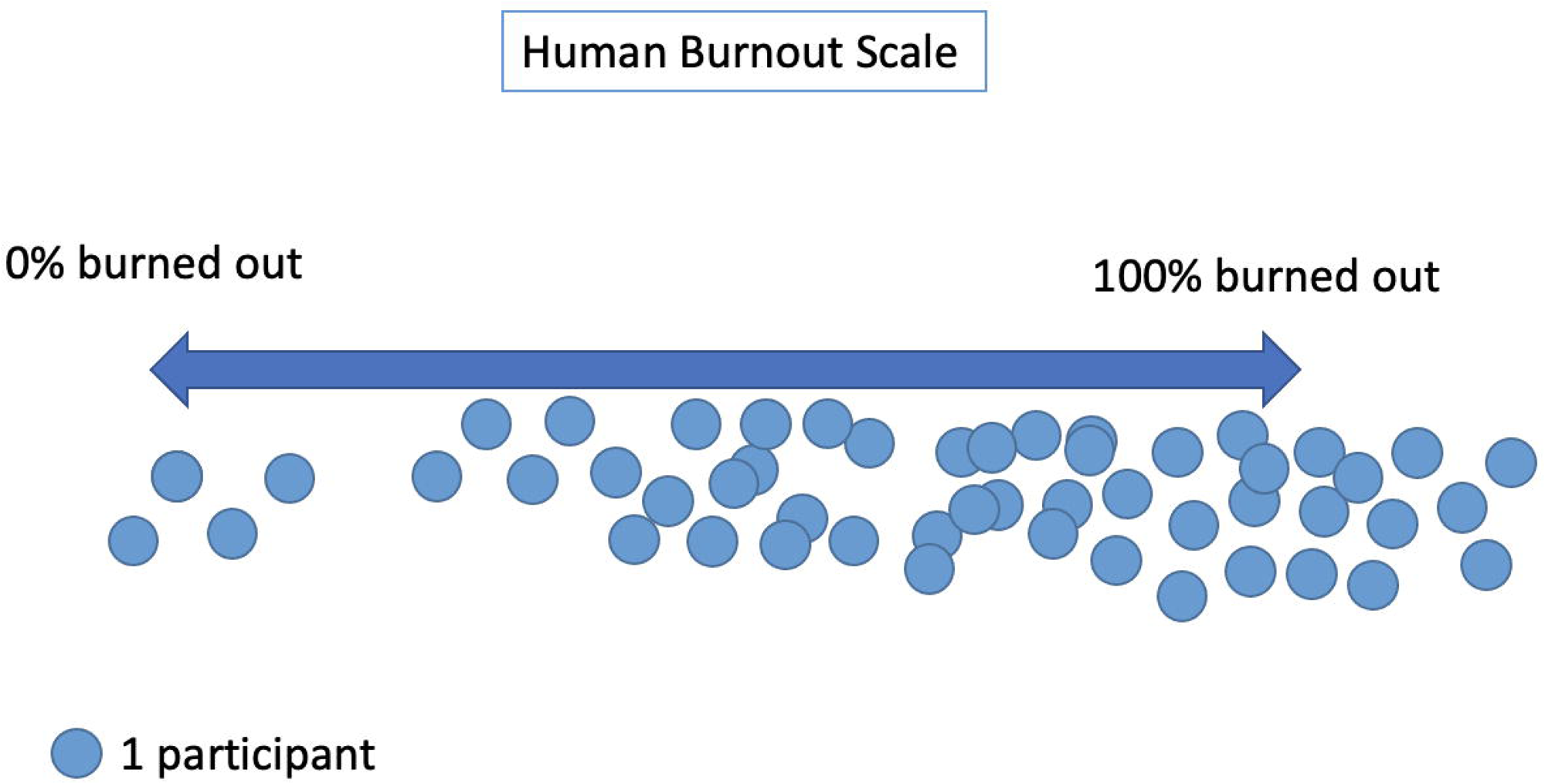
Graphic representation of participants’ placement along the human scale, combining Trainings 1 & 2.

### Assessment of burnout over time

Survey data showed that the percentage of providers meeting criteria for burnout significantly varied across the study timepoints (p<0.001), as seen in Figure 2. Immediately after the discussion of burnout and interactive activity, burnout was reassessed, and prevalence rose from 11 (18%) at baseline to 37 (62%), a number closer to what was seen in the human scale created during the activity. One month post training, burnout decreased significantly from the mid-training exercise (62% vs. 29%, p<0.001), with the impact sustained (33%) at 3 months post training (Figure 2).

**Figure 2:**
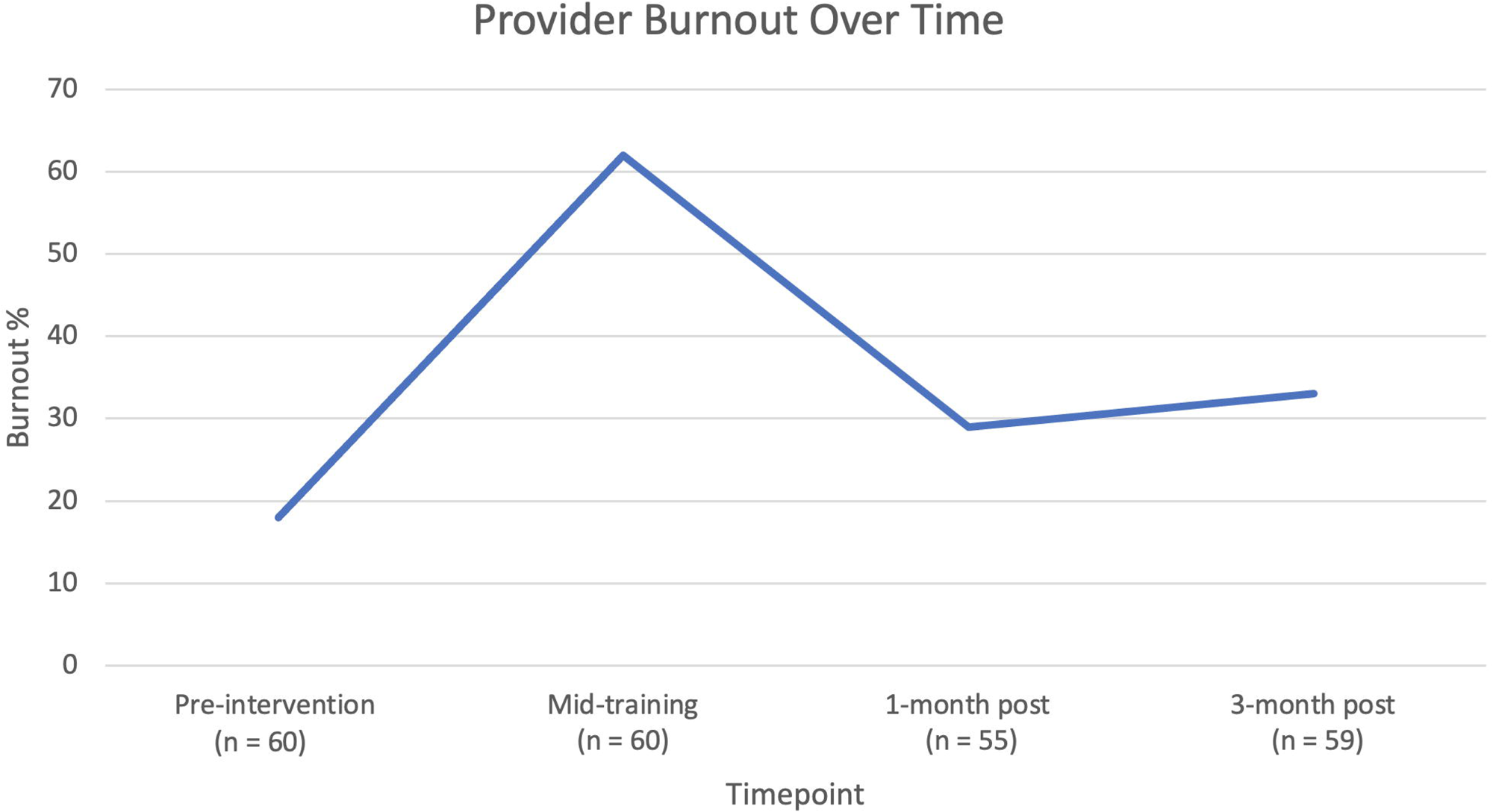
Change in proportion of providers meeting burnout criteria over time

### In-depth interviews

The IDIs with 15 L&D providers (refer to Table 1 for a description of the interviewees) offered additional insight into the experience of burnout in this population and an explanation of the change in burnout over time. The majority of respondents (14/15) believed that burnout was widespread in their workplace. Table 2 describes the themes that emerged relating to prevalence of burnout, drivers of burnout, manifestations of burnout, explanations for low baseline burnout scores, and explanation of changes in reported burnout over time.

**Table 2:**
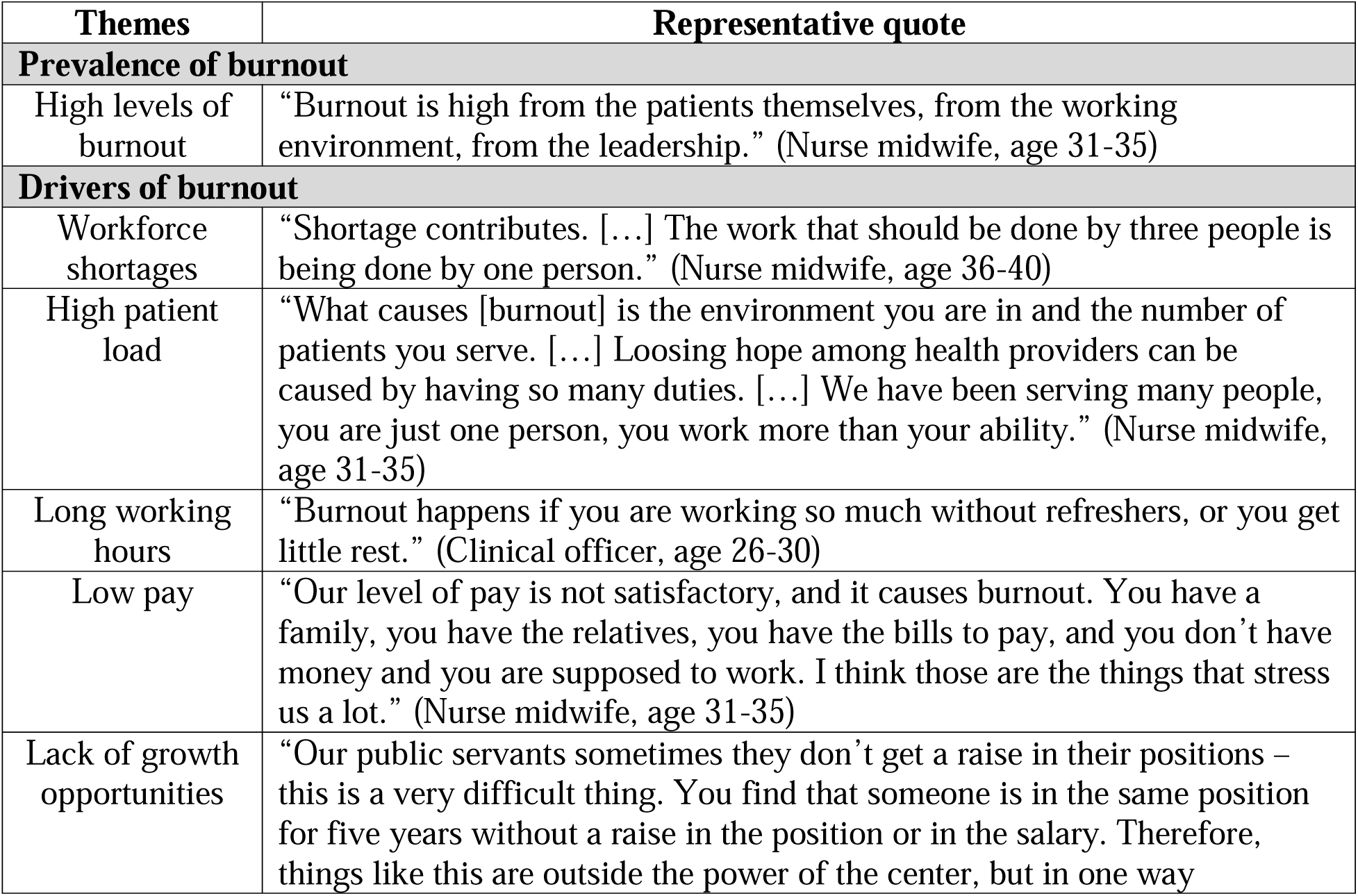
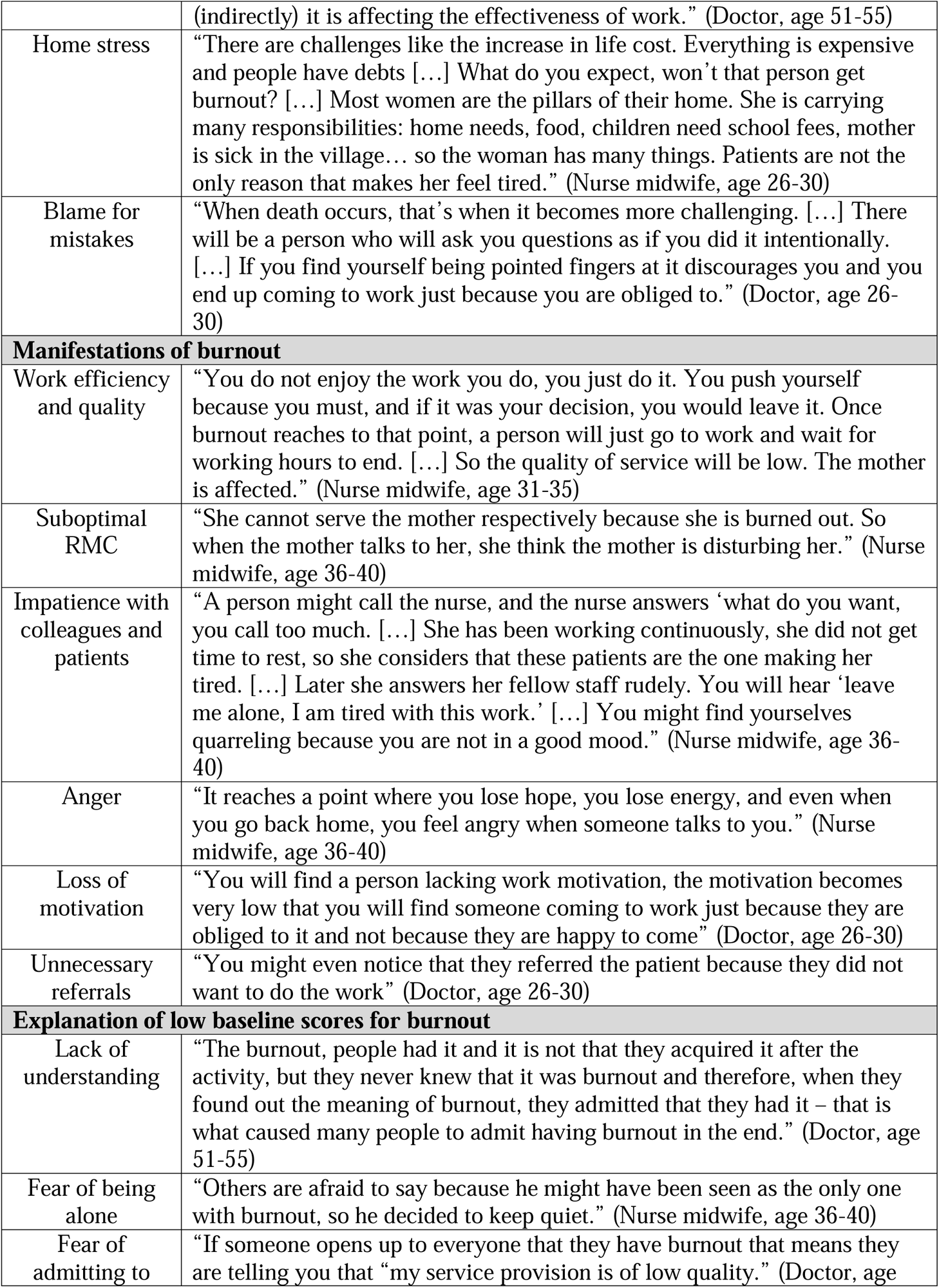
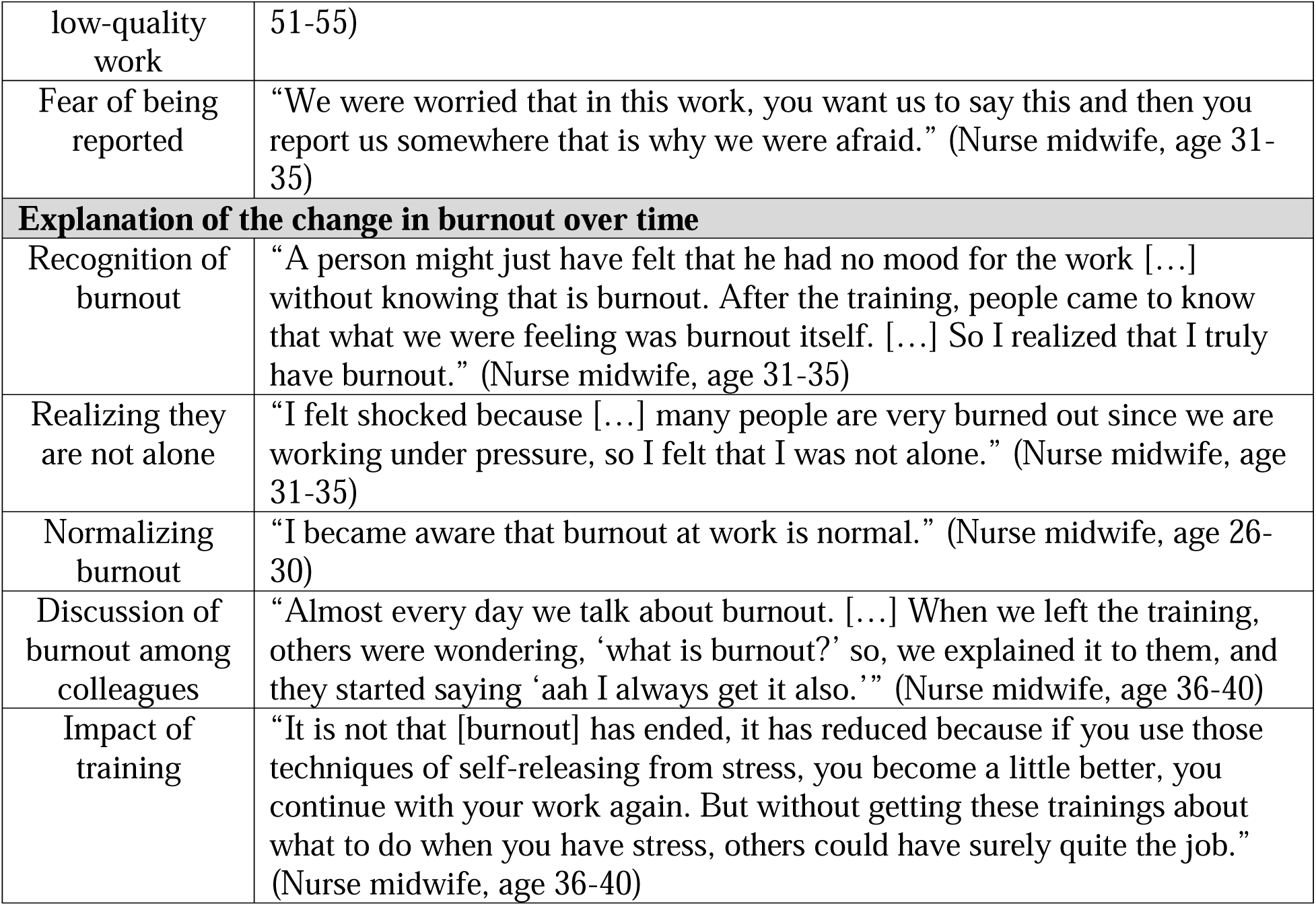
In-depth interview emerging themes

### Drivers of burnout

In discussing drivers of burnout in their clinical setting, respondents identified workforce shortages and high patient load as the most common contributors, along with low pay, long working hours, limited resources, and lack of opportunities for professional growth. Participants spoke about how clinical factors led to an inability to provide quality patient care. Several participants also discussed home stress and pressure from leadership as contributors to burnout.

### Manifestations of burnout

When asked how burnout manifests in the clinical setting, most respondents discussed the impact of burnout on work efficiency and quality. Other manifestations of burnout that emerged included suboptimal respectful maternity care, impatience both with colleagues and patients, loss of motivation in one’s work, and anger. One interviewee pointed to an increased number of unnecessary referrals.

### Explanation for low baseline scores

When asked why baseline (pre-activity) burnout scores might have been low despite experiences of burnout among providers, participants all responded that this was due to a lack of understanding of the word burnout and unfamiliarity with the concept. A few interviewees also suggested respondents may have felt reluctant to share at first due to a fear of being the only one experiencing burnout, a feeling they would be admitting to low quality work, or a fear of being reported to leadership by the study team.

### Explanation of changes in reported burnout over time

In discussing the impact of the training and activity on the providers’ experience of burnout, a majority of respondents said the activity enabled providers to recognize burnout as an issue, to realize they were not alone experiencing the feelings of burnout, and to normalize burnout as a common issue among their colleagues, leading to an increase in burnout levels reported immediately after the training activity. The training also generated further discussion around burnout among providers upon returning to work, and all interviewees emphasized how useful the coping strategies discussed during the training have been in helping them manage their stress. They all attributed the decrease in burnout prevalence seen at one- and three-months post training to the impact of the training and use of the skills they acquired and practiced there.

## DISCUSSION

Burnout among healthcare workers in Sub-Saharan Africa remains underexplored and poorly understood. However, its impact on provider wellbeing, patient outcomes and healthcare systems is undeniable. In this paper, we present the findings from our study of provider burnout among L&D providers in Tanzania, explored through survey assessments, a team activity, and in-depth interviews. Results speak to the need for better assessment of burnout for better monitoring and evaluation and for identifying intervention approaches to address this issue.

### Measurement of burnout

To understand the burden of burnout, we must first be able to identify and measure it. And if burnout is not understood, existing measures cannot be reliably applied. The survey we used was based on a validated two-item measure of burnout (20). However, at baseline before the training session on burnout, most providers scored low on the burnout scale, with 18% meeting criteria for burnout, which was not consistent with our clinical observations and previous focus group discussions with providers. As we began the discussion of burnout during the training, we realized that most participants had not previously heard of either the term or concept of burnout, and that it is not a topic commonly discussed in Tanzania. This was confirmed during our in-depth interviews (IDIs), in which providers all expressed that prior to the training, burnout was not a concept they were familiar with, despite experiencing the symptoms of burnout. The use of the activity to define and discuss burnout gave the providers a “name” for feelings that they were already experiencing.

Following the discussion and activity, the participants’ scores on the repeated burnout scale were significantly higher, with 62% meeting criteria for burnout. When discussing this change during the IDIs, providers said this rise in reported burnout represented a new understanding of the term and a recognition of pre-existing symptoms as burnout. This showed us that the validated burnout measures cannot be implemented in a new setting without first explaining and discussing burnout. If providers are not familiar with the concept, the survey will not accurately represent the level of burnout in their clinical setting.

### Addressing burnout

Providers commonly identified workforce shortages, high patient load, low pay, long hours, and limited resources as causes of burnout, which are recurring themes in previous literature on burnout in Sub-Saharan Africa and clearly not modifiable through provider training (12,15) However, one critical beginning step in addressing burnout is enabling providers to recognize it as an issue. The interactive activity showed that a majority of providers were experiencing symptoms of burnout, and this helped providers see that they were not alone in their feelings of exhaustion and depersonalization due to their job, but that this was an experience shared by many of their colleagues. This was a very important step in recognizing the issue as collective and systemic. This process demonstrates the importance of identifying burnout as an issue and encouraging discussion among providers in order for it to be recognized and addressed. Providers reported during the IDIs that, as a result of this activity, burnout has become a commonly discussed issue in the workplace. These findings are consistent with previous research, which has shown that sharing personal experiences with peers reduces professional isolation (26) and that interventions promoting collegiality, shared experience, and community among healthcare providers can improve meaning, engagement and empowerment at work (27).

Second, providers need strategies to mitigate the impact of burnout on their wellbeing and patient care. Through the training, we discussed mindfulness strategies that participants could implement immediately upon returning to work to help them manage their stress on a day-to-day basis. During the in-depth interviews, we learned that these coping strategies have had a large impact on providers’ experience of burnout and have created camaraderie as they help each other implement the various strategies to cope with stress. The survey data one- and three-months post-training revealed relatively low burnout rates again, at 29% and 33%, respectively, showing that the impact of the coping methods on reported levels of burnout endures three months after the training. As expressed by one of our interviewees, while the environment has not changed, these numbers show that training participants now have the tools to manage burnout and therefore feel less burdened by it daily.

Finally, institutional approaches alongside individual strategies are necessary to promote provider well-being (27). While these individual behavioral coping methods may be helpful in the short term, broader systems changes are needed to address the core causes of burnout among providers, requiring structural changes and increased funding of the public healthcare system.

### Limitations

This study engaged labor and delivery providers across a range of clinical settings (public and faith-based) in one region of Tanzania, making the results generalizable to a range of settings in Tanzania and perhaps beyond. The results, however, must be interpreted in the light of the study limitations. One limitation is the length of the quantitative burnout assessment. While the two-item measure has been validated previously (19), it remains a more limited evaluation of burnout compared to the full 22-item Maslach Burnout Inventory. However, the ability to rapidly administer the survey was necessary in our setting, enabling us to collect data at four time points, and the two-item measure holds a balanced part within our mixed methods. One major limitation is the absence of a comparison condition to conclusively evaluate the impact of the intervention on provider burnout. However, the IDIs enabled us to get a deeper understanding of the training impact and pointed to an improvement in provider burnout as a result of the training content. Finally, due to the small sample size, we were not able to stratify data by demographics or professional characteristics.

## CONCLUSION

Burnout is a global issue with harmful impacts on healthcare provider health, patient outcomes, and health system function. In low-resource settings, burnout is exacerbated by workforce shortages, high patient loads, and limited resources. Yet the prevalence and impact of burnout among healthcare providers in SSA has rarely been studied. This study sheds light on the experience of burnout in Tanzania and can help provide guidance on future assessments of burnout and interventions in this setting. Increasing awareness of the issue is crucial to enable providers to recognize burnout, and further investment in individual and system-wide strategies to reduce burnout are needed.

## Funding

This research is funded by the Fogarty International Center in the U.S. National Institutes of Health (R21 TW012001). Virginie Marchand is funded by a training award from the Office of AIDS Research and Fogarty International Center in the U.S. National Institutes of Health (D43 TW010543).

## Data Availability

Data is available as supplemental material.

## Acknowledgements

We are grateful for the support of the Kilimanjaro Regional Medical Office, the leadership of participating clinics, and all participating providers for their time, input, and the quality of care they provide to their patients.

## Supplemental material

**S1.** Consort diagram

**S2.** Raw data from quantitative provider surveys at four timepoints

## Notes

### Competing Interest Statement

The authors have declared no competing interest.

### Clinical Trial

NCT05271903

### Funding Statement

The study is funded by the National Institutes of Health (R21 TW012001) and by the Office of AIDS Research and Fogarty International Center in the U.S. National Institutes of Health (D43 TW010543) and is registered on clinicaltrials.gov (NCT05271903).

### Author Declarations

The study was approved by the ethical review committees at the University of Utah (Protocol 00143918), Kilimanjaro Christian Medical Center (Protocol 2056), and National Institute for Medical Research in Tanzania (Protocol 3853).

## REFERENCES

1. Chutiyami M, Cheong AMY, Salihu D, Bello UM, Ndwiga D, Maharaj R, et al. COVID-19 Pandemic and Overall Mental Health of Healthcare Professionals Globally: A Meta-Review of Systematic Reviews. Front Psychiatry. 2022 Jan 17;12:804525.

2. Meira-Silva VST, Freire ACTN, Zinezzi DP, Ribeiro FCR, Coutinho GD, Lima IMB, et al. Burnout syndrome in healthcare workers during the COVID-19 pandemic: a systematic review. Rev Bras Med Trab. 20(1):122–31.

3. West CP, Dyrbye LN, Shanafelt TD. Physician burnout: contributors, consequences and solutions. J Intern Med. 2018;283(6):516–29.

4. Dyrbye LN, West CP, Satele D, Boone S, Tan L, Sloan J, et al. Burnout Among U.S. Medical Students, Residents, and Early Career Physicians Relative to the General U.S. Population. Acad Med. 2014 Mar;89(3):443.

5. Bianchi R, Schonfeld IS, Laurent E. Burnout–depression overlap: A review. Clin Psychol Rev. 2015 Mar 1;36:28–41.

6. Oreskovich MR, Kaups KL, Balch CM, Hanks JB, Satele D, Sloan J, et al. Prevalence of alcohol use disorders among American surgeons. Arch Surg Chic Ill 1960. 2012 Feb;147(2):168–74.

7. van der Heijden F, Dillingh G, Bakker A, Prins J. Suicidal thoughts among medical residents with burnout. Arch Suicide Res Off J Int Acad Suicide Res. 2008;12(4):344–6.

8. Shanafelt TD, Balch CM, Bechamps G, Russell T, Dyrbye L, Satele D, et al. Burnout and Medical Errors Among American Surgeons. Ann Surg. 2010 Jun;251(6):995.

9. Dewa CS, Loong D, Bonato S, Thanh NX, Jacobs P. How does burnout affect physician productivity? A systematic literature review. BMC Health Serv Res. 2014 Jul 28;14:325.

10. Opoku DA, Ayisi-Boateng NK, Osarfo J, Sulemana A, Mohammed A, Spangenberg K, et al. Attrition of Nursing Professionals in Ghana: An Effect of Burnout on Intention to Quit. Nurs Res Pract. 2022;2022:3100344.

11. Robbins R, Butler M, Schoenthaler A. Provider burnout and patient-provider communication in the context of hypertension care. Patient Educ Couns. 2019 Aug 1;102(8):1452–9.

12. Dubale BW, Friedman LE, Chemali Z, Denninger JW, Mehta DH, Alem A, et al. Systematic review of burnout among healthcare providers in sub-Saharan Africa. BMC Public Health. 2019 Sep 11;19:1247.

13. Dugani S, Afari H, Hirschhorn LR, Ratcliffe H, Veillard J, Martin G, et al. Prevalence and factors associated with burnout among frontline priMary health care providers in low- and middle-income countries: A systematic review. Gates Open Res. 2018 Jun 11;2:4.

14. van der Doef M, Mbazzi FB, Verhoeven C. Job conditions, job satisfaction, somatic complaints and burnout among East African nurses. J Clin Nurs. 2012;21(11–12):1763–75.

15. Makuku R, Mosadeghrad AM. Health workforce retention in low-income settings: an application of the Root Stem Model. J Public Health Policy. 2022;43(3):445–55.

16. Anyangwe SCE, Mtonga C. Inequities in the Global Health Workforce: The Greatest Impediment to Health in Sub-Saharan Africa. Int J Environ Res Public Health. 2007 Feb;4(2):93–100.

17. Thorsen VC, Tharp ALT, Meguid T. High rates of burnout among maternal health staff at a referral hospital in Malawi: A cross-sectional study. BMC Nurs. 2011 May 23;10:9.

18. Watt MH, Minja LM, Barabara M, Mlay P, Stephens MJ, Olomi G, et al. A simulation and experiential learning intervention for labor and delivery providers to address HIV stigma during childbirth in Tanzania: study protocol for the evaluation of the MAMA intervention. BMC Pregnancy Childbirth. 2023 Mar 16;23(1):181.

19. West CP, Dyrbye LN, Sloan JA, Shanafelt TD. Single Item Measures of Emotional Exhaustion and Depersonalization Are Useful for Assessing Burnout in Medical Professionals. J Gen Intern Med. 2009 Dec;24(12):1318–21.

20. Maslach C, Leiter MP. Understanding the burnout experience: recent research and its implications for psychiatry. World Psychiatry. 2016;15(2):103–11.

21. Dolan ED, Mohr D, Lempa M, Joos S, Fihn SD, Nelson KM, et al. Using a Single Item to Measure Burnout in Primary Care Staff: A Psychometric Evaluation. J Gen Intern Med. 2015 May;30(5):582–7.

22. Morse J. Critical Issues in Qualitative Research Methods. Thousand Oaks, CA: SAGE Publications; 1994.

23. Guest G, MacQueen K, Namey E. Applied Thematic Analysis. Thousand Oaks, CA: SAGE Publications, Inc.; 2012.

24. Strauss A, Corbin J. Basics of qualitative research (3rd edition): Techniques and procedures for developing grounded theory. Thousand Oaks, California: SAGE Publications, Inc.; 2012.

25. Saunders B, Sim J, Kingstone T, Baker S, Waterfield J, Bartlam B, et al. Saturation in qualitative research: exploring its conceptualization and operationalization. Qual Quant. 2018;52(4):1893–907.

26. Beckman HB, Wendland M, Mooney C, Krasner MS, Quill TE, Suchman AL, et al. The impact of a program in mindful communication on primary care physicians. Acad Med J Assoc Am Med Coll. 2012 Jun;87(6):815–9.

27. West CP, Dyrbye LN, Rabatin JT, Call TG, Davidson JH, Multari A, et al. Intervention to promote physician well-being, job satisfaction, and professionalism: a randomized clinical trial. JAMA Intern Med. 2014 Apr;174(4):527–33.

