## Supplemental figure 1 for "Burnout among labor and birth providers in northern Tanzania: A mixed-method study": S1 Figure 1. Consort diagram.docx

Supplemental table 1. Consort diagram

**Invited to participate**

**(n=60)**

Changed jobs (n=1)

Maternity leave (n=1)

Busy with clinical duties (n=2)

Loss to follow up (n=1)

**Attended *In Situ* training**

**(n=55)**

**Completed Immediate Post Assessment**

**(n=60)**

**Attended MAMA training**

**(n=60)**

**Completed 3 Month Assessment**

**(n=59)**

**Completed 1 Month Assessment**

**(n=55)**

**Completed Baseline Assessment**

**(n=60)**

**Enrolled**

**(n=60)**
