## Supplemental data for "Burnout among labor and birth providers in northern Tanzania: A mixed-method study"

| ID | timepoint | dem1 | dem2 | dem3 | dem3_other | dem4 | dem5 | dem6 | dem7 | burn1 | burn2 |
|---|---|---|---|---|---|---|---|---|---|---|---|
| 1 | pre-training | 47 | 1 | 3 | NA | 20 | 15 | 1 | 1 | 1 | 1 |
| 1 | mid-training | NA | NA | NA | NA | NA | NA | NA | NA | 1 | 2 |
| 1 | 1 month-training | NA | NA | NA | NA | NA | NA | NA | NA | 1 | 1 |
| 1 | 3 month-training | NA | NA | NA | NA | NA | NA | NA | NA | 1 | 1 |
| 2 | pre-training | 28 | 1 | 2 | NA | 7 | 5 | 5 | 1 | 1 | 0 |
| 2 | mid-training | NA | NA | NA | NA | NA | NA | NA | NA | 2 | 1 |
| 2 | 1 month-training | NA | NA | NA | NA | NA | NA | NA | NA | 2 | 0 |
| 2 | 3 month-training | NA | NA | NA | NA | NA | NA | NA | NA | 2 | 0 |
| 3 | pre-training | 33 | 1 | 2 | NA | 10 | 4 | 7 | 4 | 0 | 3 |
| 3 | mid-training | NA | NA | NA | NA | NA | NA | NA | NA | 2 | 5 |
| 3 | 1 month-training | NA | NA | NA | NA | NA | NA | NA | NA | 3 | 5 |
| 3 | 3 month-training | NA | NA | NA | NA | NA | NA | NA | NA | 3 | 5 |
| 4 | pre-training | 39 | 1 | 4 | NA | 14 | 6 | 0.3 | 1 | 4 | 1 |
| 4 | mid-training | NA | NA | NA | NA | NA | NA | NA | NA | 3 | 0 |
| 4 | 1 month-training | NA | NA | NA | NA | NA | NA | NA | NA | NA | NA |
| 4 | 3 month-training | NA | NA | NA | NA | NA | NA | NA | NA | 0 | 0 |
| 5 | pre-training | 28 | 1 | 2 | NA | 1 | 1 | 1 | 1 | 0 | 1 |
| 5 | mid-training | NA | NA | NA | NA | NA | NA | NA | NA | 3 | 1 |
| 5 | 1 month-training | NA | NA | NA | NA | NA | NA | NA | NA | NA | NA |
| 5 | 3 month-training | NA | NA | NA | NA | NA | NA | NA | NA | 0 | 1 |
| 6 | pre-training | 28 | 1 | 2 | NA | 7 | 1 | 2 | 1 | 4 | 1 |
| 6 | mid-training | NA | NA | NA | NA | NA | NA | NA | NA | 4 | 1 |
| 6 | 1 month-training | NA | NA | NA | NA | NA | NA | NA | NA | 4 | 1 |
| 6 | 3 month-training | NA | NA | NA | NA | NA | NA | NA | NA | 0 | 1 |
| 7 | pre-training | 29 | 1 | 2 | NA | 6 | 1 | NA | 1 | 4 | 1 |
| 7 | mid-training | NA | NA | NA | NA | NA | NA | NA | NA | 4 | 1 |
| 7 | 1 month-training | NA | NA | NA | NA | NA | NA | NA | NA | 0 | 1 |
| 7 | 3 month-training | NA | NA | NA | NA | NA | NA | NA | NA | 0 | 0 |
| 8 | pre-training | 28 | 2 | 2 | NA | 4 | 3 | 3 | 2 | 0 | 0 |
| 8 | mid-training | NA | NA | NA | NA | NA | NA | NA | NA | 2 | 3 |
| 8 | 1 month-training | NA | NA | NA | NA | NA | NA | NA | NA | 0 | 1 |
| 8 | 3 month-training | NA | NA | NA | NA | NA | NA | NA | NA | 4 | 1 |
| 9 | pre-training | 49 | 1 | 2 | NA | 29 | 20 | 6 | 1 | 1 | 2 |
| 9 | mid-training | NA | NA | NA | NA | NA | NA | NA | NA | 1 | 1 |
| 9 | 1 month-training | NA | NA | NA | NA | NA | NA | NA | NA | 1 | 0 |
| 9 | 3 month-training | NA | NA | NA | NA | NA | NA | NA | NA | 0 | 1 |
| 10 | pre-training | 33 | 1 | 4 | NA | 2 | 2 | 2 | 2 | 1 | 2 |
| 10 | mid-training | NA | NA | NA | NA | NA | NA | NA | NA | 1 | 3 |
| 10 | 1 month-training | NA | NA | NA | NA | NA | NA | NA | NA | 0 | 0 |
| 10 | 3 month-training | NA | NA | NA | NA | NA | NA | NA | NA | 0 | 0 |
| 11 | pre-training | 52 | 2 | 4 | NA | 20 | 20 | 3 | 1 | 0 | 0 |
| 11 | mid-training | NA | NA | NA | NA | NA | NA | NA | NA | 0 | 0 |
| 11 | 1 month-training | NA | NA | NA | NA | NA | NA | NA | NA | 1 | 1 |
| 11 | 3 month-training | NA | NA | NA | NA | NA | NA | NA | NA | 1 | 0 |
| 12 | pre-training | 32 | 2 | 2 | NA | 8 | 5 | 7 | 1 | 0 | 0 |
| 12 | mid-training | NA | NA | NA | NA | NA | NA | NA | NA | 1 | 0 |
| 12 | 1 month-training | NA | NA | NA | NA | NA | NA | NA | NA | 0 | 0 |
| 12 | 3 month-training | NA | NA | NA | NA | NA | NA | NA | NA | 0 | 0 |
| 13 | pre-training | 30 | 2 | 2 | NA | 3 | 2 | 1 | 1 | 0 | 0 |
| 13 | mid-training | NA | NA | NA | NA | NA | NA | NA | NA | 3 | 2 |
| 13 | 1 month-training | NA | NA | NA | NA | NA | NA | NA | NA | 1 | 2 |
| 13 | 3 month-training | NA | NA | NA | NA | NA | NA | NA | NA | 0 | 2 |
| 14 | pre-training | 27 | 2 | 2 | NA | 3 | 0.3 | 0.3 | 1 | 0 | 0 |
| 14 | mid-training | NA | NA | NA | NA | NA | NA | NA | NA | 0 | 0 |
| 14 | 1 month-training | NA | NA | NA | NA | NA | NA | NA | NA | 0 | 0 |
| 14 | 3 month-training | NA | NA | NA | NA | NA | NA | NA | NA | 0 | 0 |
| 15 | pre-training | 39 | 1 | 4 | NA | 5 | 5 | 5 | 1 | 1 | 1 |
| 15 | mid-training | NA | NA | NA | NA | NA | NA | NA | NA | 2 | 1 |
| 15 | 1 month-training | NA | NA | NA | NA | NA | NA | NA | NA | 0 | 1 |
| 15 | 3 month-training | NA | NA | NA | NA | NA | NA | NA | NA | 0 | 1 |
| 16 | pre-training | 53 | 2 | 4 | NA | 25 | 12 | 12 | 1 | 2 | 5 |
| 16 | mid-training | NA | NA | NA | NA | NA | NA | NA | NA | 2 | 3 |
| 16 | 1 month-training | NA | NA | NA | NA | NA | NA | NA | NA | 0 | 3 |
| 16 | 3 month-training | NA | NA | NA | NA | NA | NA | NA | NA | 0 | 1 |
| 17 | pre-training | 34 | 1 | 2 | NA | 12 | 4 | 12 | 1 | 2 | 1 |
| 17 | mid-training | NA | NA | NA | NA | NA | NA | NA | NA | 2 | 1 |
| 17 | 1 month-training | NA | NA | NA | NA | NA | NA | NA | NA | 2 | 3 |

| | | | | | | | | | | | |
|---|---|---|---|---|---|---|---|---|---|---|---|
| 17 | 3 month-training | NA | NA | NA | NA | NA | NA | NA | NA | 1 | 0 |
| 18 | pre-training | 28 | 1 | 2 | NA | 6 | 1 | 1 | 1 | 2 | 3 |
| 18 | mid-training | NA | NA | NA | NA | NA | NA | NA | NA | 2 | 3 |
| 18 | 1 month-training | NA | NA | NA | NA | NA | NA | NA | NA | 1 | 3 |
| 18 | 3 month-training | NA | NA | NA | NA | NA | NA | NA | NA | 1 | 5 |
| 19 | pre-training | 34 | 2 | 2 | NA | 11 | 6 | 10 | 2 | 0 | 1 |
| 19 | mid-training | NA | NA | NA | NA | NA | NA | NA | NA | 2 | 3 |
| 19 | 1 month-training | NA | NA | NA | NA | NA | NA | NA | NA | 1 | 2 |
| 19 | 3 month-training | NA | NA | NA | NA | NA | NA | NA | NA | 1 | 1 |
| 20 | pre-training | 52 | 2 | 4 | NA | 28 | 10 | 4 | 1 | 1 | 1 |
| 20 | mid-training | NA | NA | NA | NA | NA | NA | NA | NA | 1 | 1 |
| 20 | 1 month-training | NA | NA | NA | NA | NA | NA | NA | NA | 0 | 1 |
| 20 | 3 month-training | NA | NA | NA | NA | NA | NA | NA | NA | 0 | 1 |
| 21 | pre-training | 32 | 1 | 2 | NA | 9 | 7 | 9 | 1 | 0 | 0 |
| 21 | mid-training | NA | NA | NA | NA | NA | NA | NA | NA | 0 | 1 |
| 21 | 1 month-training | NA | NA | NA | NA | NA | NA | NA | NA | 0 | 1 |
| 21 | 3 month-training | NA | NA | NA | NA | NA | NA | NA | NA | 0 | 0 |
| 22 | pre-training | 25 | 2 | 2 | NA | 3 | 2 | 2 | 2 | 0 | 1 |
| 22 | mid-training | NA | NA | NA | NA | NA | NA | NA | NA | 1 | 2 |
| 22 | 1 month-training | NA | NA | NA | NA | NA | NA | NA | NA | 0 | 1 |
| 22 | 3 month-training | NA | NA | NA | NA | NA | NA | NA | NA | 1 | 1 |
| 23 | pre-training | 26 | 2 | 3 | NA | 1 | 1 | 1 | 1 | 1 | 1 |
| 23 | mid-training | NA | NA | NA | NA | NA | NA | NA | NA | 2 | 1 |
| 23 | 1 month-training | NA | NA | NA | NA | NA | NA | NA | NA | 2 | 1 |
| 23 | 3 month-training | NA | NA | NA | NA | NA | NA | NA | NA | 2 | 1 |
| 24 | pre-training | 27 | 2 | 4 | NA | 2 | 2 | 1 | 1 | 1 | 0 |
| 24 | mid-training | NA | NA | NA | NA | NA | NA | NA | NA | 2 | 1 |
| 24 | 1 month-training | NA | NA | NA | NA | NA | NA | NA | NA | 1 | 1 |
| 24 | 3 month-training | NA | NA | NA | NA | NA | NA | NA | NA | 1 | 1 |
| 25 | pre-training | NA | 2 | 2 | NA | 2 | 2 | 1 | 1 | 0 | 0 |
| 25 | mid-training | NA | NA | NA | NA | NA | NA | NA | NA | 2 | 1 |
| 25 | 1 month-training | NA | NA | NA | NA | NA | NA | NA | NA | NA | NA |
| 25 | 3 month-training | NA | NA | NA | NA | NA | NA | NA | NA | 4 | 1 |
| 26 | pre-training | 32 | 1 | 2 | NA | 10 | 10 | 9 | 1 | 0 | 0 |
| 26 | mid-training | NA | NA | NA | NA | NA | NA | NA | NA | 3 | 1 |
| 26 | 1 month-training | NA | NA | NA | NA | NA | NA | NA | NA | 3 | 1 |
| 26 | 3 month-training | NA | NA | NA | NA | NA | NA | NA | NA | 4 | 1 |
| 27 | pre-training | 30 | 2 | 2 | NA | 10 | 3 | 9 | 1 | 0 | 0 |
| 27 | mid-training | NA | NA | NA | NA | NA | NA | NA | NA | 2 | 1 |
| 27 | 1 month-training | NA | NA | NA | NA | NA | NA | NA | NA | 2 | 0 |
| 27 | 3 month-training | NA | NA | NA | NA | NA | NA | NA | NA | 1 | 1 |
| 28 | pre-training | 52 | 2 | 4 | NA | 17 | 12 | 12 | 1 | 0 | 0 |
| 28 | mid-training | NA | NA | NA | NA | NA | NA | NA | NA | 0 | 0 |
| 28 | 1 month-training | NA | NA | NA | NA | NA | NA | NA | NA | 0 | 0 |
| 28 | 3 month-training | NA | NA | NA | NA | NA | NA | NA | NA | 0 | 0 |
| 29 | pre-training | 29 | 2 | 4 | NA | 2 | 1 | 1 | 1 | 3 | 2 |
| 29 | mid-training | NA | NA | NA | NA | NA | NA | NA | NA | 3 | 3 |
| 29 | 1 month-training | NA | NA | NA | NA | NA | NA | NA | NA | 3 | 3 |
| 29 | 3 month-training | NA | NA | NA | NA | NA | NA | NA | NA | 4 | 3 |
| 30 | pre-training | 45 | 1 | 2 | NA | 18 | 10 | 10 | 1 | 0 | 3 |
| 30 | mid-training | NA | NA | NA | NA | NA | NA | NA | NA | 1 | 0 |
| 30 | 1 month-training | NA | NA | NA | NA | NA | NA | NA | NA | 0 | 0 |
| 30 | 3 month-training | NA | NA | NA | NA | NA | NA | NA | NA | 0 | 0 |
| 31 | pre-training | 32 | 2 | 3 | NA | 8 | 8 | 8 | 1 | 0 | 0 |
| 31 | mid-training | NA | NA | NA | NA | NA | NA | NA | NA | 3 | 3 |
| 31 | 1 month-training | NA | NA | NA | NA | NA | NA | NA | NA | 1 | 1 |
| 31 | 3 month-training | NA | NA | NA | NA | NA | NA | NA | NA | 1 | 1 |
| 32 | pre-training | 37 | 2 | 2 | NA | 8 | 8 | 8 | 2 | 1 | 2 |
| 32 | mid-training | NA | NA | NA | NA | NA | NA | NA | NA | 4 | 3 |
| 32 | 1 month-training | NA | NA | NA | NA | NA | NA | NA | NA | 1 | 1 |
| 32 | 3 month-training | NA | NA | NA | NA | NA | NA | NA | NA | 1 | 1 |
| 33 | pre-training | 28 | 2 | 2 | NA | 6 | 2 | 1 | 3 | 0 | 0 |
| 33 | mid-training | NA | NA | NA | NA | NA | NA | NA | NA | 2 | 0 |
| 33 | 1 month-training | NA | NA | NA | NA | NA | NA | NA | NA | 1 | 1 |
| 33 | 3 month-training | NA | NA | NA | NA | NA | NA | NA | NA | 3 | 0 |
| 34 | pre-training | 34 | 1 | 2 | NA | 11 | 11 | 11 | 1 | 0 | 0 |
| 34 | mid-training | NA | NA | NA | NA | NA | NA | NA | NA | 2 | 5 |
| 34 | 1 month-training | NA | NA | NA | NA | NA | NA | NA | NA | 2 | 0 |

| | | | | | | | | | | | |
|---|---|---|---|---|---|---|---|---|---|---|---|
| 34 | 3 month-training | NA | NA | NA | NA | NA | NA | NA | NA | 2 | 0 |
| 35 | pre-training | 29 | 2 | 4 | NA | 3 | 1 | 1 | 2 | 0 | 1 |
| 35 | mid-training | NA | NA | NA | NA | NA | NA | NA | NA | 3 | 5 |
| 35 | 1 month-training | NA | NA | NA | NA | NA | NA | NA | NA | 1 | 3 |
| 35 | 3 month-training | NA | NA | NA | NA | NA | NA | NA | NA | 1 | 1 |
| 36 | pre-training | 43 | 2 | 4 | NA | 20 | 10 | 1 | 4 | 0 | 1 |
| 36 | mid-training | NA | NA | NA | NA | NA | NA | NA | NA | 1 | 1 |
| 36 | 1 month-training | NA | NA | NA | NA | NA | NA | NA | NA | 2 | 3 |
| 36 | 3 month-training | NA | NA | NA | NA | NA | NA | NA | NA | 2 | 3 |
| 37 | pre-training | 26 | 1 | 2 | NA | 6 | 4 | 1 | 2 | 0 | 2 |
| 37 | mid-training | NA | NA | NA | NA | NA | NA | NA | NA | 3 | 3 |
| 37 | 1 month-training | NA | NA | NA | NA | NA | NA | NA | NA | NA | NA |
| 37 | 3 month-training | NA | NA | NA | NA | NA | NA | NA | NA | 1 | 4 |
| 38 | pre-training | 30 | 2 | 3 | NA | 6 | 4 | 4 | 2 | 1 | 2 |
| 38 | mid-training | NA | NA | NA | NA | NA | NA | NA | NA | 3 | 4 |
| 38 | 1 month-training | NA | NA | NA | NA | NA | NA | NA | NA | 2 | 1 |
| 38 | 3 month-training | NA | NA | NA | NA | NA | NA | NA | NA | 0 | 1 |
| 39 | pre-training | 34 | 2 | 2 | NA | 8 | 7 | 7 | 1 | 4 | 5 |
| 39 | mid-training | NA | NA | NA | NA | NA | NA | NA | NA | 2 | 5 |
| 39 | 1 month-training | NA | NA | NA | NA | NA | NA | NA | NA | 1 | 3 |
| 39 | 3 month-training | NA | NA | NA | NA | NA | NA | NA | NA | 1 | 2 |
| 40 | pre-training | 30 | 1 | 2 | NA | 10 | 9 | 9 | 2 | 2 | 6 |
| 40 | mid-training | NA | NA | NA | NA | NA | NA | NA | NA | 2 | 5 |
| 40 | 1 month-training | NA | NA | NA | NA | NA | NA | NA | NA | 2 | 1 |
| 40 | 3 month-training | NA | NA | NA | NA | NA | NA | NA | NA | 3 | 3 |
| 41 | pre-training | 35 | 2 | 4 | NA | 2 | 2 | 1 | 1 | 1 | 3 |
| 41 | mid-training | NA | NA | NA | NA | NA | NA | NA | NA | 3 | 5 |
| 41 | 1 month-training | NA | NA | NA | NA | NA | NA | NA | NA | 1 | 3 |
| 41 | 3 month-training | NA | NA | NA | NA | NA | NA | NA | NA | 1 | 2 |
| 42 | pre-training | 32 | 2 | 2 | NA | 5 | 4 | 5 | 1 | 1 | 1 |
| 42 | mid-training | NA | NA | NA | NA | NA | NA | NA | NA | 3 | 2 |
| 42 | 1 month-training | NA | NA | NA | NA | NA | NA | NA | NA | 1 | 0 |
| 42 | 3 month-training | NA | NA | NA | NA | NA | NA | NA | NA | 1 | 0 |
| 43 | pre-training | 29 | 1 | 2 | NA | 8 | 6 | 2 | 1 | 1 | 1 |
| 43 | mid-training | NA | NA | NA | NA | NA | NA | NA | NA | 1 | 1 |
| 43 | 1 month-training | NA | NA | NA | NA | NA | NA | NA | NA | 1 | 1 |
| 43 | 3 month-training | NA | NA | NA | NA | NA | NA | NA | NA | 0 | 1 |
| 44 | pre-training | 32 | 2 | 2 | NA | 6 | 3 | 1 | 1 | 1 | 1 |
| 44 | mid-training | NA | NA | NA | NA | NA | NA | NA | NA | 2 | 1 |
| 44 | 1 month-training | NA | NA | NA | NA | NA | NA | NA | NA | 0 | 0 |
| 44 | 3 month-training | NA | NA | NA | NA | NA | NA | NA | NA | 2 | 1 |
| 45 | pre-training | 37 | 1 | 2 | NA | 13 | 7 | 8 | 1 | 0 | 0 |
| 45 | mid-training | NA | NA | NA | NA | NA | NA | NA | NA | 1 | 1 |
| 45 | 1 month-training | NA | NA | NA | NA | NA | NA | NA | NA | 0 | 0 |
| 45 | 3 month-training | NA | NA | NA | NA | NA | NA | NA | NA | 0 | 2 |
| 46 | pre-training | 25 | 2 | 2 | NA | 4 | 2 | 1 | 2 | 0 | 2 |
| 46 | mid-training | NA | NA | NA | NA | NA | NA | NA | NA | 4 | 4 |
| 46 | 1 month-training | NA | NA | NA | NA | NA | NA | NA | NA | 2 | 2 |
| 46 | 3 month-training | NA | NA | NA | NA | NA | NA | NA | NA | 2 | 2 |
| 47 | pre-training | 46 | 2 | 2 | NA | 11 | 11 | 11 | 1 | 4 | 3 |
| 47 | mid-training | NA | NA | NA | NA | NA | NA | NA | NA | 1 | 1 |
| 47 | 1 month-training | NA | NA | NA | NA | NA | NA | NA | NA | 0 | 1 |
| 47 | 3 month-training | NA | NA | NA | NA | NA | NA | NA | NA | 0 | 0 |
| 48 | pre-training | 42 | 2 | 4 | NA | 14 | 7 | 14 | 1 | 0 | 0 |
| 48 | mid-training | NA | NA | NA | NA | NA | NA | NA | NA | 0 | 0 |
| 48 | 1 month-training | NA | NA | NA | NA | NA | NA | NA | NA | 0 | 0 |
| 48 | 3 month-training | NA | NA | NA | NA | NA | NA | NA | NA | 0 | 0 |
| 49 | pre-training | 31 | 2 | 4 | NA | 2 | 2 | 2 | 1 | 0 | 0 |
| 49 | mid-training | NA | NA | NA | NA | NA | NA | NA | NA | 0 | 0 |
| 49 | 1 month-training | NA | NA | NA | NA | NA | NA | NA | NA | NA | NA |
| 49 | 3 month-training | NA | NA | NA | NA | NA | NA | NA | NA | NA | NA |
| 50 | pre-training | 29 | 1 | 1 | NA | 9 | 5 | 9 | 2 | 0 | 0 |
| 50 | mid-training | NA | NA | NA | NA | NA | NA | NA | NA | 0 | 0 |
| 50 | 1 month-training | NA | NA | NA | NA | NA | NA | NA | NA | 0 | 0 |
| 50 | 3 month-training | NA | NA | NA | NA | NA | NA | NA | NA | 0 | 0 |
| 51 | pre-training | 35 | 1 | 2 | NA | 8 | 6 | 8 | 1 | 1 | 1 |
| 51 | mid-training | NA | NA | NA | NA | NA | NA | NA | NA | 3 | 1 |
| 51 | 1 month-training | NA | NA | NA | NA | NA | NA | NA | NA | 1 | 1 |

| | | | | | | | | | | | |
|---|---|---|---|---|---|---|---|---|---|---|---|
| 51 | 3 month-training | NA | NA | NA | NA | NA | NA | NA | NA | 1 | 1 |
| 52 | pre-training | 36 | 1 | 2 | NA | 11 | 10 | 11 | 1 | 1 | 1 |
| 52 | mid-training | NA | NA | NA | NA | NA | NA | NA | NA | 1 | 1 |
| 52 | 1 month-training | NA | NA | NA | NA | NA | NA | NA | NA | 0 | 1 |
| 52 | 3 month-training | NA | NA | NA | NA | NA | NA | NA | NA | NA | NA |
| 53 | pre-training | NA | 1 | 2 | NA | 30 | 10 | 30 | 1 | 0 | 0 |
| 53 | mid-training | NA | NA | NA | NA | NA | NA | NA | NA | 2 | 1 |
| 53 | 1 month-training | NA | NA | NA | NA | NA | NA | NA | NA | 0 | 3 |
| 53 | 3 month-training | NA | NA | NA | NA | NA | NA | NA | NA | 0 | 1 |
| 54 | pre-training | 40 | 1 | 2 | NA | 8 | 4 | 5 | 1 | 4 | 0 |
| 54 | mid-training | NA | NA | NA | NA | NA | NA | NA | NA | 3 | 1 |
| 54 | 1 month-training | NA | NA | NA | NA | NA | NA | NA | NA | 3 | 1 |
| 54 | 3 month-training | NA | NA | NA | NA | NA | NA | NA | NA | 3 | 1 |
| 55 | pre-training | 29 | 2 | 4 | NA | 3 | 0.25 | 0.25 | 1 | 0 | 0 |
| 55 | mid-training | NA | NA | NA | NA | NA | NA | NA | NA | 1 | 0 |
| 55 | 1 month-training | NA | NA | NA | NA | NA | NA | NA | NA | 2 | 0 |
| 55 | 3 month-training | NA | NA | NA | NA | NA | NA | NA | NA | 2 | 0 |
| 56 | pre-training | 57 | 2 | 4 | NA | 10 | 6 | 6 | 1 | 0 | 2 |
| 56 | mid-training | NA | NA | NA | NA | NA | NA | NA | NA | 2 | 2 |
| 56 | 1 month-training | NA | NA | NA | NA | NA | NA | NA | NA | 0 | 1 |
| 56 | 3 month-training | NA | NA | NA | NA | NA | NA | NA | NA | 2 | 1 |
| 57 | pre-training | 40 | 1 | 2 | NA | 15 | 7 | 15 | 1 | 0 | 0 |
| 57 | mid-training | NA | NA | NA | NA | NA | NA | NA | NA | 1 | 5 |
| 57 | 1 month-training | NA | NA | NA | NA | NA | NA | NA | NA | 2 | 3 |
| 57 | 3 month-training | NA | NA | NA | NA | NA | NA | NA | NA | 4 | 1 |
| 58 | pre-training | 55 | 1 | 2 | NA | 25 | 8 | 25 | 1 | 0 | 0 |
| 58 | mid-training | NA | NA | NA | NA | NA | NA | NA | NA | 0 | 0 |
| 58 | 1 month-training | NA | NA | NA | NA | NA | NA | NA | NA | 0 | 0 |
| 58 | 3 month-training | NA | NA | NA | NA | NA | NA | NA | NA | 0 | 0 |
| 59 | pre-training | 28 | 1 | 2 | NA | 9 | 4 | 4 | 1 | 0 | 0 |
| 59 | mid-training | NA | NA | NA | NA | NA | NA | NA | NA | 0 | 0 |
| 59 | 1 month-training | NA | NA | NA | NA | NA | NA | NA | NA | 0 | 0 |
| 59 | 3 month-training | NA | NA | NA | NA | NA | NA | NA | NA | 0 | 0 |
| 60 | pre-training | 38 | 1 | 2 | NA | 8 | 4 | 2 | 1 | 0 | 0 |
| 60 | mid-training | NA | NA | NA | NA | NA | NA | NA | NA | 1 | 2 |
| 60 | 1 month-training | NA | NA | NA | NA | NA | NA | NA | NA | 0 | 0 |
| 60 | 3 month-training | NA | NA | NA | NA | NA | NA | NA | NA | 0 | 0 |

dem1 How old are you?
dem2 What is your gender [1=Female, 2=Male]
dem3 What is your clinical training [1=Midwife, 2=Nurse midwife, 3=Clinical officer, 4=Medical doctor, 5=Other]
dem4 How many years has it been since you completed your clinical schooling?
dem5 How many years have you been providing care in labor and delivery?
dem6 How many years have you been working at the clinic/hospital where youre currently employed?
dem7 In the last year, how many trainings have you completed that included content about labor and delivery for women with HIV
burn1 Using your own definition of burnout, please select one of the following responses [0= I enjoy my work. I have no symptoms of burnout. 1= Occasionally I am under stress, and I don't always have as much energy as I once did, but I don't feel burned out. 2= I am definitely burning out and have one or more symptoms of burnout, such as physical and emotional exhaustion. 3= The symptoms of burnout I am experiencing won't go away. I think about frustrations at work a lot. 5= I feel completely burned out and often wonder if I can go on. I am at the point where I may need some changes or may need to seek some sort of help.]
burn2 How often do you feel youve become more callous toward people since you took this job because of burnout? [0= Never, 1= A few times a year or less, 2= Once a month or less, 3= A few times a month, 4= Once a week, 5= A few times a week, 6= Every day]
